# Preterm Preeclampsia Risk Modelling: Examining Hemodynamic, Biochemical, and Biophysical Markers Prior to Pregnancy

**DOI:** 10.1101/2023.02.28.23286590

**Authors:** Bryn C. Loftness, Ira Bernstein, Carole A. McBride, Nick Cheney, Ellen W. McGinnis, Ryan S. McGinnis

## Abstract

Preeclampsia (PE) is a leading cause of maternal and perinatal death globally and can lead to unplanned preterm birth. Predicting risk for preterm or early-onset PE, has been investigated primarily after conception, and particularly in the early and mid-gestational periods. However, there is a distinct clinical advantage in identifying individuals at risk for PE prior to conception, when a wider array of preventive interventions are available. In this work, we leverage machine learning techniques to identify potential pre-pregnancy biomarkers of PE in a sample of 80 women, 10 of whom were diagnosed with preterm preeclampsia during their subsequent pregnancy. We explore biomarkers derived from hemodynamic, biophysical, and biochemical measurements and several modeling approaches. A support vector machine (SVM) optimized with stochastic gradient descent yields the highest overall performance with ROC AUC and detection rates up to .88 and .70, respectively on subject-wise cross validation. The best performing models leverage biophysical and hemodynamic biomarkers. While preliminary, these results indicate the promise of a machine learning based approach for detecting individuals who are at risk for developing preterm PE before they become pregnant. These efforts may inform gestational planning and care, reducing risk for adverse PE-related outcomes.

**Clinical Relevance:** This work considers the development and optimization of pre-pregnancy biomarkers for improving the identification of preterm (early-onset) preeclampsia risk prior to conception.

## I. Introduction

Preeclampsia (PE) is a hypertensive disorder that affects up to 5% of pregnancies and is associated with elevated maternal and perinatal mortality [1,2]. In recent years, work on predictive modeling of PE risk during gestation has emerged [2,3], but few studies consider data prior to pregnancy [4,5]. Identifying the pathological underpinnings of PE distinguishable pre-pregnancy could enable screening tools for prospective child-bearers, the deployment of proactive care plans, and novel interventions, reducing the threat of maternal and perinatal morbidity and mortality during gestation and post-birth. Past work has laid the groundwork in identifying and classifying preeclamptic phenotypes of differing risk levels during gestation [6,7,8]. These investigations have allowed for new interventions to be tested and validated to reduce risk of preterm PE, starting in the first trimester [9]. Efforts towards identifying phenotypes of preterm PE risk could have meaningful impacts, as those with preterm PE have been found to have significantly higher extended risk of mortality than child bearers who did not have PE or individuals with PE who were able to carry to term [10]–[12].

Previous work has begun to validate the combination of biophysical and biochemical testing as integrative components for advancing predictive modeling of PE early in pregnancy [6,13,14]. Our past work has identified significant differences in the hemodynamics of individuals with prior preeclampsia *vs* individuals without a prior pregnancy and presented preliminary importance of pre-pregnancy features to the subsequent development of preterm complicated hypertension [15,16]. This study complements past work by continuing to assess individuals with PE prior to pregnancy leveraging new machine learning techniques. We collect modalities shown to be strong predictors in previous efforts, as well as examine additional candidate features that may be unique to pre-gestational monitoring— addressing the need for models dedicated to pre-pregnancy risk screening for preterm PE.

Mirroring the incremental biomarker addition methodology in [13,17,18], we leverage machine learning techniques to examine features derived from baseline maternal cardiovascular hemodynamics (HD), biochemical (via blood test) biomarkers (BC), and biophysical cardiac measurements (BP) for detecting risk for preterm PE prior to pregnancy. These evaluations point to the potential for deploying devices and procedures to screen for preterm PE risk prior to pregnancy, allowing for enhanced proactive care and planning.

## II. Methods

This is a secondary analysis of a prospective longitudinal study in which women were recruited prior to planned pregnancy and followed for pregnancy outcomes. Following delivery, medical records were reviewed for diagnosis of preeclampsia. At the time of study, subjects were healthy, non-smoking, and normotensive. Subjects were recruited through open advertisement and were compensated for their time. Out of an original 124 subjects in the study, data from 80 subjects were considered for analysis based on adequate amounts of data collected. During their subsequent pregnancy, 10 (12.5%) were diagnosed with preterm PE, four (5%) with PE at term, and 13 (16.25%) with gestational hypertension. Fifty subjects were nulliparous, 27 had a history of preterm preeclampsia in their most recent pregnancy, three did not have a history of preeclampsia in their recent pregnancy. Pre-pregnancy assessments were made during the follicular phase of the menstrual cycle, on mean cycle day 9±4. Assessments were made following a three-day sodium/potassium-controlled diet and in a post-absorptive fasting state. Subjects were 31±4 years (mean ± S.D.) and had mean body mass index (BMI) of 25.6±5.6 kg/m^2^.

Hemodynamic (HD) data were collected continuously via a Finipres Pro (FMS, Netherlands) with non-invasive tonometric assessment. The HD data included a one-minute baseline period in a rested supine position. Four subjects had less than 60 seconds of valid baseline data (24, 42, 48, and 54 seconds), but were still included in the 80 total subjects considered. HD data included beat-by-beat heart rate, systolic blood pressure, diastolic blood pressure, pulse interval, cardiac output, mean arterial pressure, stroke volume, left ventricular ejection time, maximum slope, and total peripheral resistance. A number of these modalities are employed for gestational biomarker measurements included within predictive models for preeclampsia and hypertensive disorders of pregnancy [6,3]. For each of these continuous data streams, the following statistical features were computed using tsfresh [19]: coefficient of variation, mean absolute changes, mean, min, max, time series complexity (CIDce), and absolute sum of changes.

Blood was collected from the clinical data visit for analyses of chemical screening (BC) to document insulin resistance (HomaIR), C-Reactive Protein, soluble amyloid precursor protein alpha (s-APPα) and s-APP beta, which have been identified as being increased in women who develop preeclampsia [20,21]. Four subjects, with normotensive pregnancy outcomes did not have biochemical testing, but were included in this study. For any model that used BC markers, we elected to remove them from the training data rather than to impute their values. Each subject also had biophysical (BP) data collected during the visit: intravascular plasma volume corrected for lean body mass, renal cortical resistance index (RI), and two measures of aortic-popliteal pulse wave velocity. Consistent with previous studies [6,3,22], maternal clinical characteristics including age and BMI were also tested for applicability within the sample. We do not assess several features common in previous work, such as first trimester assessment of uterine hemodynamics or early pregnancy serum markers of placental function as the focus of this work is the pre-pregnancy period.

We evaluated performance of three classifiers deployed with shuffled 2-fold cross validation for predicting preterm PE— gaussian naïve Bayes (NB), support vector machine optimized using stochastic gradient descent (SVM), and random forest (RF)— chosen largely based on past work within PE predictive modelling [6,3]. We present data from an analysis of seven modality combinations, each tested on the three classifiers: BP, BC, HD, HD + BP, HD + BC, BP + BC, and HD + BC + BP. With each combination, we also consider maternal BMI and age.

Of these classifiers and modality combinations, we identify the top models based on each of the performance metrics: ROC AUC (presented as mean of the test sets), false positive rate, and true positive rate, consistent with past literature [6]. We then identify the top performing classifier across the spectrum of our analysis and present the performance metrics based on the various modality-based feature combinations for that classifier. We conclude with a breakdown of top-ranking features from each modality after feature reduction, wherein candidate features were selected via univariate analysis with a significance level of .05, mirroring the methodology of several works reviewed in [6].

## III. Results

### A. Top Performing Models and Modalities

The top 5 highest performing models based on each of the performance metrics are presented in **Table 1**. Based upon maximization of ROC AUC and true positive rate, the models built with the SVM had the highest performance. Compared to statistical surveys for the relevance of maternal screening guidelines such as those provided by National Institute for Health and Care Excellence (NICE) and the American College of Obstetricians and Gynecologists (ACOG), our models perform on-par with the published detection rates, which are based largely on individual and family diagnostic history and maternal health characteristics. Between these two organizations, they present detection rates of 5-41%, with a .2-10% false positive rate [9]. Our top models produced 50-70% detection rates, with 3-16% false positive rate. This indicates that data collected pre-pregnancy has the potential to classify risk of preterm preeclampsia with similar or superior performance to current nationally accepted and widely used screening procedures.

**Table 1.**
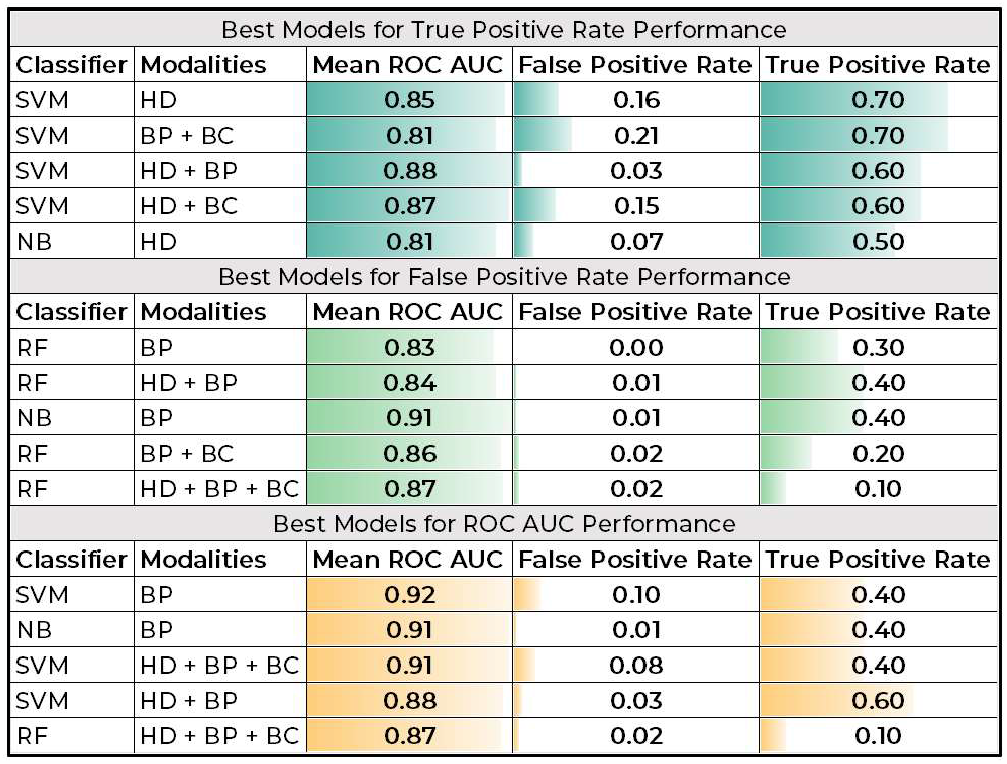
Model Performance by Classifier and Modality Integration Scenario. Mean ROC AUC across k-fold test groups, as well as overall false positive rate and true positive rate from prediction across k-fold groups grouped by classifier and modality scenario. Only the top five highest performing models based on each of the three metrics are shown.

| Best Models for True Positive Rate Performance |  |  |  |  |
| --- | --- | --- | --- | --- |
| Classifier | Modalities | Mean ROC AUC | False Positive Rate | True Positive Rate |
| SVM | HD | 0.85 | 0.16 | 0.70 |
| SVM | BP + BC | 0.81 | 0.21 | 0.70 |
| SVM | HD + BP | 0.88 | 0.03 | 0.60 |
| SVM | HD + BC | 0.87 | 0.15 | 0.60 |
| NB | HD | 0.81 | 0.07 | 0.50 |
| Best Models for False Positive Rate Performance |  |  |  |  |
| Classifier | Modalities | Mean ROC AUC | False Positive Rate | True Positive Rate |
| RF | BP | 0.83 | 0.00 | 0.30 |
| RF | HD + BP | 0.84 | 0.01 | 0.40 |
| NB | BP | 0.91 | 0.01 | 0.40 |
| RF | BP + BC | 0.86 | 0.02 | 0.20 |
| RF | HD + BP + BC | 0.87 | 0.02 | 0.10 |
| Best Models for ROC AUC Performance |  |  |  |  |
| Classifier | Modalities | Mean ROC AUC | False Positive Rate | True Positive Rate |
| SVM | BP | 0.92 | 0.10 | 0.40 |
| NB | BP | 0.91 | 0.01 | 0.40 |
| SVM | HD + BP + BC | 0.91 | 0.08 | 0.40 |
| SVM | HD + BP | 0.88 | 0.03 | 0.60 |
| RF | HD + BP + BC | 0.87 | 0.02 | 0.10 |

Model performance (ROC AUCs between .8-.92) is in line with results from a robust and modern review of prediction models for PE based on data collected *during* pregnancy [6]. The review featured studies with ROC AUCs ranging .65-.98. Considering our dataset only includes data recorded pre-pregnancy at varying time windows to conception, the resulting model performance is promising.

Examining results from the best performing SVM model more closely, **Figure 1** highlights the relative performance of each modality combination. Across modalities, ROC AUC varied only slightly between the top 6 modality combinations (∼.8-.92), with the two highest performing models considering only biophysical data with maternal BMI and a model combining all three modalities with maternal BMI. We also observe relatively low false positive rates (<∼.2) across all modalities. This result, in combination with the fact that six out of seven of the SVM model assessments’ true positive rates are greater than or equal to 0.4, bodes well for a relatively wide, but accurate net being cast for identifying preterm preeclamptic risk across models using the SVM classifier.

**Figure 1.**
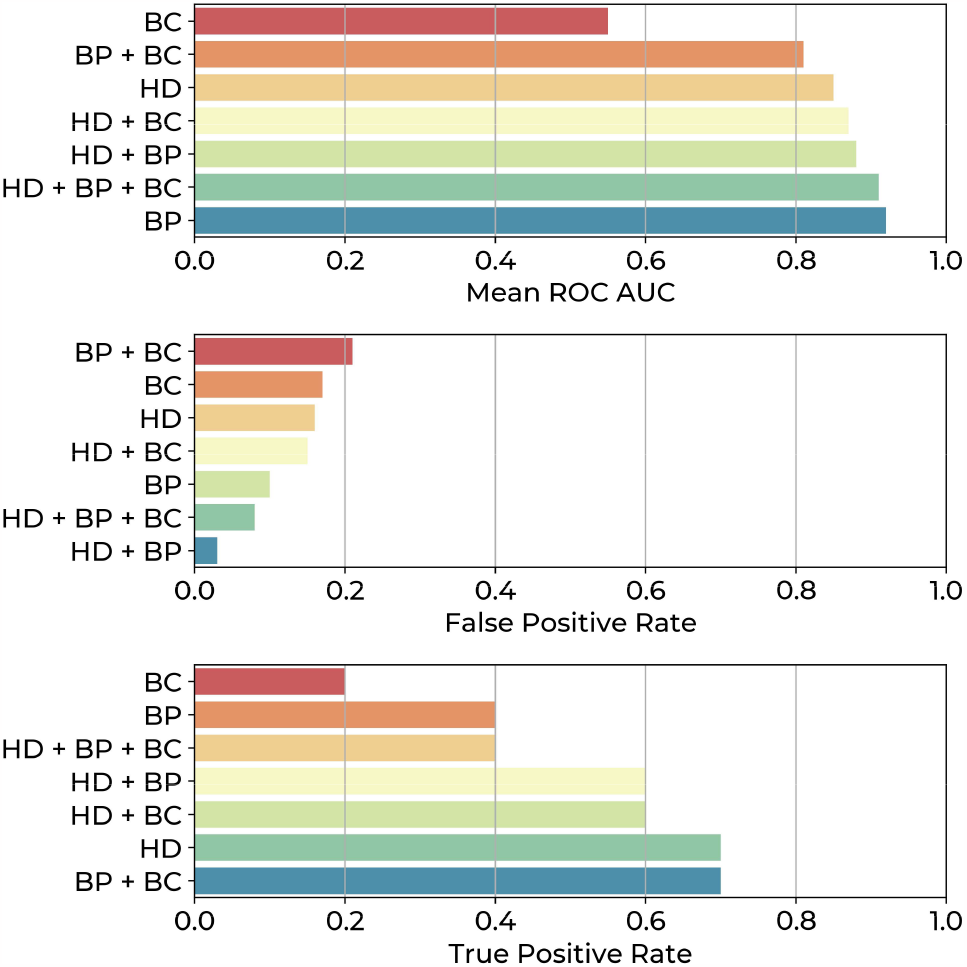
Model Performance by Modality Integration Scenario for only SVM Models. Mean ROC AUC, overall false positive rates, and true positive rates from test-case prediction across k-fold groups aggregated by modality scenario.

### B. Feature Reduction and Diagnostic-Specific Differences

For each modality, univariate significance testing yielded a substantial reduction in the number of features (**Table 2**). Of the 83 initial features from the HD, BP, BC, and maternal characteristics data sources, only 24 features remained. Sixteen of those features were HD variables, 4 were BP, and 3 were BC. Of the maternal characteristics of age and BMI, only BMI was significant and thus was included in model testing. Several biomarker trends emerging through these feature reduction efforts are consistent with international guidelines on maternal biomarkers for preeclampsia risk. Specifically, we observed increased systolic and diastolic blood pressure, and higher average BMI, which provides additional support for these results [5,9,23].

**Table 2.**
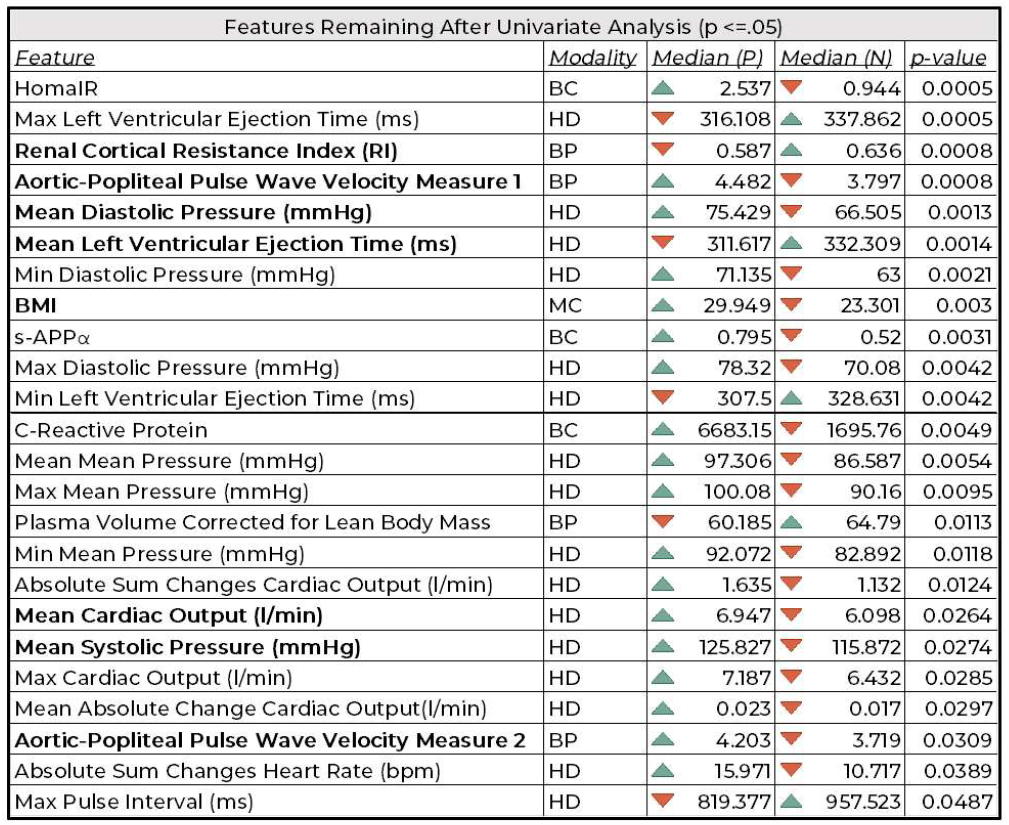
Features Remaining After Univariate Significance Analysis. Each feature with a significant group-level difference between preterm preeclamptic individuals (P) versus all others in the sample population (N), evaluated via Mann Whitney U. Modality, Medians, and p-values are reported for each feature. Arrows indicate the higher and lower median across the two groups for each feature. Top-ranking biomarkers based on permutation testing are bolded.

Permutation analysis of each model suggests that models including the BP features of aortic-popliteal pulse wave velocity and renal cortical resistance index consistently ranked those features as being highly influential in model prediction. Maternal BMI was also consistently regarded as influential for model performance across the modality combinations. For BC and HD features, the top-ranking biomarkers are more varied but often pertain to mean systolic pressure and diastolic pressure, mean left ventricular ejection time, and cardiac output. Further testing must be completed to establish more robust feature importance rankings within and across modalities.

## IV. Discussion

In this study we explore machine learning based approaches for detecting women, pre-pregnancy, who will develop preterm PE. Data collected from a sample of 80 adult women pre-pregnancy were used to explore three classification models and seven combinations of measurement modalities to identify two best performing model-modality combinations. These models achieved both high classification performance (0.88 and .85) and detection rates (.6 and .7). Results are in line with prior work, but uniquely considers data from this pre-gestational timepoint which has significant clinical implications for managing gestational planning and care.

Notably, the models we consider are binary classifiers for identifying preterm preeclampsia, which in this study categorizes individuals in our sample with no preeclampsia diagnosis alongside individuals with diagnoses of hypertension and term preeclampsia. Further analysis of model misclassifications reveals that false positives are highly saturated by individuals of different preeclamptic diagnoses rather than no diagnosis, indicating potential biomarker similarities between preterm preeclampsia and other preeclamptic phenotypes at the pre-gestational phase. Additional data collection is needed to increase sample size of all the PE subgroups, and additional analysis is needed to develop high performing multiclass models which are sensitive to the finer-grained phenotypic differences between PE diagnoses and can stratify pre-gestational risk accordingly. Despite these misclassifications, performance of the predictive models is still impressive, and particularly considering the use of data sampled from women before they become pregnant.

Building on prior work which has examined performance of predictive models trained on features extracted based on gestational period [5], we have identified new and reoccurring candidate biomarkers pre-pregnancy which is a relatively unexplored data collection window. Interestingly, the feature reduction analysis (**Table 2**) revealed a number (16) of HD features that captured important differences between the preterm PE group and all others. Of the identified HD features, 50% captured the extremes (e.g., maximum left ventricle ejection time, minimum diastolic pressure) of the underlying cardiovascular hemodynamics timeseries which mirrors findings in other populations [24]. Given the importance of these extremes, it may follow that additional data collection activities extending beyond a simple supine baseline collection, such as collection of a subset of hemodynamic measures via wearables during daily life, may reveal hemodynamic measures more sensitive to risk for developing preterm PE and improve model performance further.

Future work recruiting additional subjects pre-pregnancy, and matched on potentially confounding factors (i.e., gravida), as well as testing additional modalities, and expanding the conditions under which data are collected will further our ability to differentiate (via modelling and biomarker analysis) preeclamptic risk groups and allowing higher confidence pre-pregnancy screening models to be developed. Successful development of these models could open the door to important advances in maternal care.

## V. Conclusion

We present results supporting the potential for pre-gestational prediction of preterm preeclampsia. Machine learning models trained on various combinations of maternal cardiovascular hemodynamics, biochemical (via blood test) biomarkers, and biophysical cardiac measurements yield results that are in line with the best-performing PE risk models in prior work but are based on novel pre-gestational data. Further development of this approach for predicting risk for developing preterm PE could have significant clinical impact, namely through increased preparation and proactive monitoring or interventions for intending child-bearers and their families.

## Data Availability

Data produced and analyzed in the present study may be available upon reasonable request to the authors.

